# Continuous genomic diversification of long polynucleotide fragments drives the emergence of new SARS-CoV-2 variants of concern

**DOI:** 10.1101/2021.12.23.21268315

**Authors:** Karthik Murugadoss, Michiel J.M. Niesen, Bharathwaj Raghunathan, Patrick J. Lenehan, Pritha Ghosh, Tyler Feener, Praveen Anand, Safak Simsek, Rohit Suratekar, Travis K. Hughes, Venky Soundararajan

**Affiliations:** nference, Cambridge, Massachusetts 02139, USA; nference, Toronto, ON M5V 1M1, Canada; nference Labs, Bengaluru, Karnataka 560017, India

## Abstract

Highly transmissible or immuno-evasive SARS-CoV-2 variants have intermittently emerged and outcompeted previously circulating strains, resulting in repeated COVID-19 surges, reinfections, and breakthrough infections in vaccinated individuals. With over 5 million SARS-CoV-2 genomes sequenced globally over the last 2 years, there is unprecedented data to decipher how competitive viral evolution results in the emergence of fitter SARS-CoV-2 variants. Much attention has been directed to studying how specific mutations in the Spike protein impact its binding to the ACE2 receptor or viral neutralization by antibodies, but there is limited knowledge of a genomic signature that is shared primarily by the sequential dominant variants. Here we introduce a methodology to quantify the genome-wide distinctiveness of polynucleotide fragments of various lengths (3-to 240-mers) that constitute SARS-CoV-2 sequences (freely available at https://academia.nferx.com/GENI). Compared to standard phylogenetic distance metrics and overall mutational load, the quantification of distinctive 9-mer polynucleotides provides a higher resolution of separation between VOCs (Reference = 89, IQR: 65-108; Alpha = 166, IQR: 150-182; Beta 130, IQR: 113-147; Gamma = 165, IQR: 152-180; Delta = 234, IQR: 216-253; and Omicron = 294, IQR: 287-315). Omicron’s exceptionally high genomic distinctiveness may confer a competitive advantage over both prior VOCs (including Delta) and the recently emerged and highly mutated B.1.640.2 (IHU) lineage. Expanding on this analysis, evaluation of genomic distinctiveness weighted by intra-lineage 9-mer conservation for 1,363 lineages annotated in GISAID highlights that genomic distinctiveness has increased over time (R2=0.37) and that VOCs score significantly higher than contemporary non-VOC lineages, with Omicron among the most distinctive lineages observed till date. This study demonstrates the value of characterizing new SARS-CoV-2 variants by their genome-wide polynucleotide distinctiveness and emphasizes the need to go beyond a narrow set of mutations at known functionally or antigenically salient sites on the Spike protein. The consistently higher distinctiveness of each emerging VOC compared to prior VOCs suggests that real-time monitoring of genomic distinctiveness would aid in more rapid assessment of viral fitness.

## Introduction

The emergence of several SARS-CoV-2 Variants of Concern (VOCs: Alpha, Beta, Gamma, Delta, Omicron) over time has resulted in repeated surges of COVID-19 cases, hospitalizations, and deaths around the globe.^1^ Phylogenetic classification in the Phylogenetic Assignment of Named Global Outbreak Lineages (PANGO) nomenclature shows that these variants have evolved from common ancestors, but none are direct descendants of one another.^2^ The PANGO lineages corresponding to these VOCs are as follows: Alpha variant (B.1.1.7 and Q lineages), Beta variant (B.1.351 and descendant lineages), Gamma variant (P.1, which is a descendant of B.1.1.28, and descendant lineages), Delta variant (B.1.617.2 and AY lineages), and Omicron variant (B.1.1.529 and BA lineages).^3^ All of these variants evolved from the B.1 lineage, while Alpha, Gamma, and Omicron share B.1.1 as an additional parent lineage. A recent non-VOC, the IHU variant (B.1.640) that was identified in France also captured interest due to the amount and nature of mutations in this variant.^4^ However, these phylogenetic classifications do not intuitively describe the degree of distinctiveness between VOCs, nor do they provide concrete insights into the genomic properties of each variant.

SARS-CoV-2, like other viruses, evolves via the introduction of mutations in its genome.^5^ In some cases, these mutations yield changes in the amino acid sequence of viral proteins. Such mutations can then be positively or negatively selected depending on their impact on various aspects of viral fitness, including transmissibility (e.g., ability to infect and/or replicate in host cells) and immune evasion (e.g., ability to avoid binding by host-derived neutralizing antibodies). It has become clear from global data sharing efforts, particularly the GISAID database (https://www.gisaid.org),^6,7^ that mutations in several regions such as the receptor binding domain (RBD) and N-terminal domain (NTD) of the Spike glycoprotein contribute to improved viral fitness.^8–13^ Significant effort has been devoted to functionally characterizing the mutations that define VOCs.^9,14–16^ However, while much attention has been paid to the consequence of individual mutations at the amino acid level, there has been limited focus on how SARS-CoV-2 evolution explores the possibilities of diversifying its “language” at the level of nucleotide sequences (i.e. towards deciphering the “words” constituting polynucleotide sequences from the viral genome). Such an understanding of the language of viral evolution could facilitate multiple dimensions of pandemic preparedness including the development and improvement of diagnostics and vaccines.^17^

We hypothesized that the emergence of more transmissible or immune evasive SARS-CoV-2 variants over time is associated with increased genomic distinctiveness from the original strain and from strains that were previously circulating. To assess this hypothesis, here we introduce a new methodology to quantify the number of distinct nucleotide n-mers (of various sizes) in VOCs to estimate the degree of viral evolution. We find that SARS-CoV-2 variants that have emerged later (Delta, B.1.640, and Omicron) indeed tend to harbor more distinct nucleotide n-mers than those that emerged earlier (Alpha, Beta, and Gamma), with the original parent strain (PANGO lineage A) expectedly having the lowest level of n-mer distinctiveness. Although correlated, this striking trend is not attributable solely to overall mutational load, nor is it explained simply by a reduction in genomic distinctiveness of early VOCs due to the emergence of later VOCs. Taken together, this study demonstrates that, over the two years of the COVID-19 pandemic (December 2019 - December 2021), SARS-CoV-2 variants with increasing nucleotide distinctiveness have evolved over time.

The concept of polynucleotide diversification introduced in this study represents a new method to map epidemiological features of SARS-CoV-2 (e.g., transmissibility and immune evasiveness) to a genomic compass. To facilitate the continued monitoring of SARS-CoV-2 evolution through this lens, we are launching a freely accessible resource for pandemic preparedness with genomic inference (“Pandemic Preparedness GENI” -- https://academia.nferx.com/GENI). We encourage further investigation to determine whether genomic distinctiveness can help to predict the likelihood that newly emerging variants will outcompete viral strains that are contemporaneously circulating in the same geographic regions.

## Results

### SARS-CoV-2 variants of concern with increasing genomic distinction evolved over time

To quantify the genomic distinctiveness of SARS-CoV-2 VOCs with respect to the original strain,^3,18^ we compared the number of distinctive nucleotide 9-mers present in the Wuhan (PANGO lineage A; original strain), Alpha, Beta, Gamma, Delta, and Omicron variants (see *Methods*, **Figure 1A**). Specifically, we performed 100,000 repetitions of an iterative sampling experiment in which we selected one SARS-CoV-2 genome corresponding to each of the variants listed above (**Table S1**). From each genome, we derived the set of unique 9-mers, and then compared these sets against each other. We determined the number of distinctive nucleotide 9-mers (DN9s) which were present in each given lineage but absent in all others. The number of DN9s appeared to correlate with time of emergence, with highest values observed for Omicron (emerged in November 2021) followed by Delta (March 2021), Alpha and Gamma (September and October 2021, respectively), and finally Beta (September 2021) (**Figures 1B-C**).

**Figure 1.**
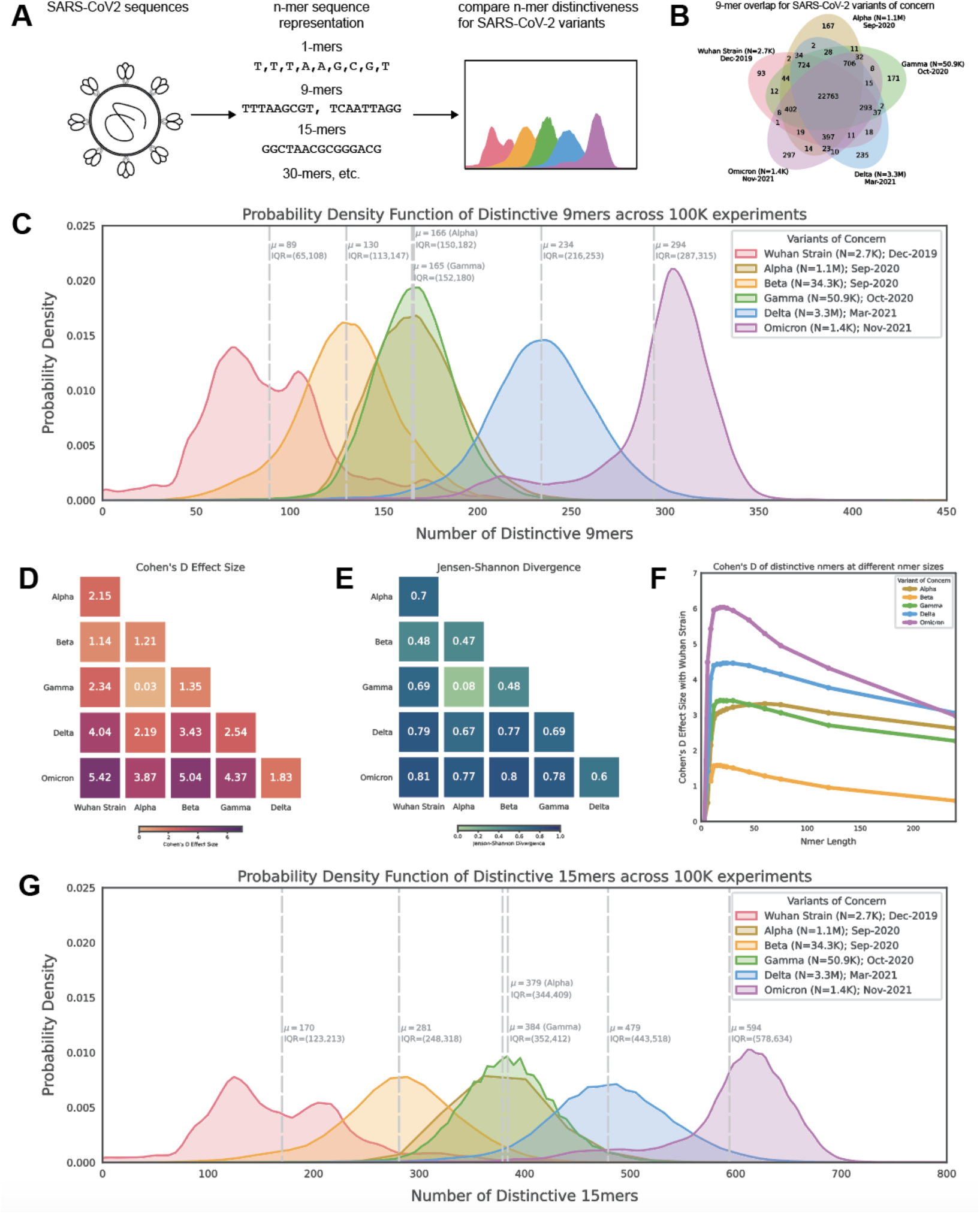
Distribution of polynucleotide distinctiveness for SARS-CoV-2 variants of concern (VOCs). **(A)** Schematic illustration of polynucleotide sequence analysis. SARS-CoV-2 sequences are analyzed to generate a set of distinct n-mer polynucleotide sequences (max n-mer size = 240). **(B)** Venn Diagram showing the mean of the distributions for shared and unique nucleotide 9-mers between all combinations of variants across 100,000 replicate comparisons. The Beta variant was excluded from this visualization to reduce clutter. **(C)** Density plots showing 9-mer sequence distinctiveness for VOCs, as measured by the number of distinct 9-mer polynucleotide sequences. **(D-E)** Heatmaps showing Cohen’s D and Jensen-Shannon divergence values from pairwise comparisons of the distributions shown in (C). **(F)** Cohen’s D of the distinctive n-mer distributions of Alpha, Beta, Gamma, Delta, and Omicron variants against the original strain for various n-mer lengths (n = 3, 6, 9, 12, 15, 18, 21, 24, 30, 45, 60, 75, 120, and 240). **(G)** Density plots showing an additional example for genomic distinctiveness of VOCs, as measured by the number of distinct 15-mer polynucleotide sequences. Data shown in panels **B-G** were generated using 287,739 unique SARS-CoV-2 sequences in total, split across the variants as shown in the legend of **C**. Abbreviations: μ - mean; IQR - interquartile range; VOC - variant of concern.

Omicron sequences robustly had more DN9s than all other VOCs (Cohen’s D > 3.0 for each comparison), and Delta sequences had more DN9s than all VOCs other than Omicron (**Figure 1C-D**). However, Alpha, Beta, and Gamma showed more overlap, with all Cohen’s D values less than 1.5 relative to each other (**Figure 1C-D**). The same pattern was also observed when comparing the DN9 distributions via Jensen-Shannon divergence (**Figure 1E**). Further, similar results were observed for various other tested lengths of polynucleotide such as 15-mers (**Figure 1F-G, Table S2**). Considering these findings, it is notable that Alpha, Beta, and Gamma emerged at similar times (September-October 2021) and drove surges in largely distinct regions (Alpha in the United States, United Kingdom, and Asia; Beta in South Africa; Gamma in South America) (**Figure S1**). On the other hand, Delta and Omicron rapidly spread across the globe and became the dominant variant in most regions after emerging in April and November 2021, respectively.^19^

### Genomic distinctiveness at the polynucleotide level provides significant additional context to existing distance metrics

We next asked whether the observed patterns in genomic distinctiveness (as captured by the DN9 metric) were captured by other simple or commonly used metrics for distinguishing SARS-CoV-2 variants. Overall mutational load does highlight Omicron as the most highly mutated VOC and is clearly correlated with our metric (**Figure 2A**). However, mutational load alone provides poor resolution between Alpha, Beta, Gamma, and Delta (**Figure 2A-C**), with mean values of 27, 26, 24, and 32, respectively. This suggests that the number of distinctive n-mers harbored by a VOC contains information beyond the number of mutations away from the original SARS-CoV-2 genome. This point is further demonstrated by the IHU variant (B.1.640.2); a sublineage of B.1.640; that emerged in October 2021 but has not spread rapidly across the globe.^4^ Despite having a higher mutational load than Delta (**Figure 3A**), these variants have similar distinctiveness at the polynucleotide level (**Figure 3B**). Together, these examples show that increasing mutational load does not imply an increase in polynucleotide distinctiveness and vice versa.

**Figure 2.**
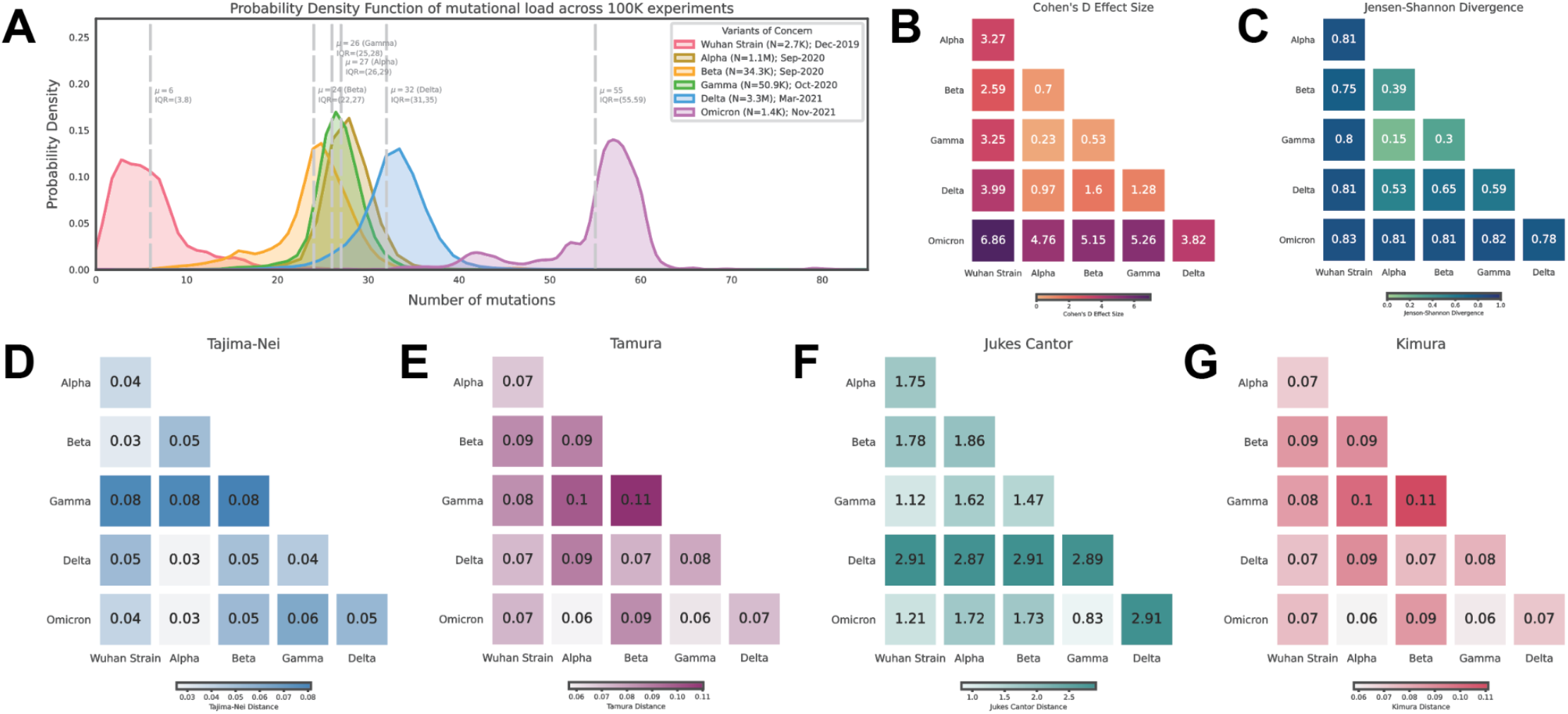
Mutational load and phylogenetic distance analysis of SARS-CoV-2 VOCs. **(A)** Density plots showing the distribution of the number of mutations observed in SARS-CoV-2 genomes assigned to each listed lineage in the GISAID database. **(B-C)** Heatmaps showing Cohen’s D and Jensen-Shannon divergence values from pairwise comparisons of the distribution of distinctive 9-mer sequences between SARS-CoV-2 VOCs. **(D-G)** Heatmaps showing the mean phylogenetic distance calculated using multiple methods (Tajima-Nei, Tamura, Jukes-Cantor, and Kimura) between SARS-CoV-2 VOCs. Abbreviations: VOC - variant of concern.

**Figure 3.**
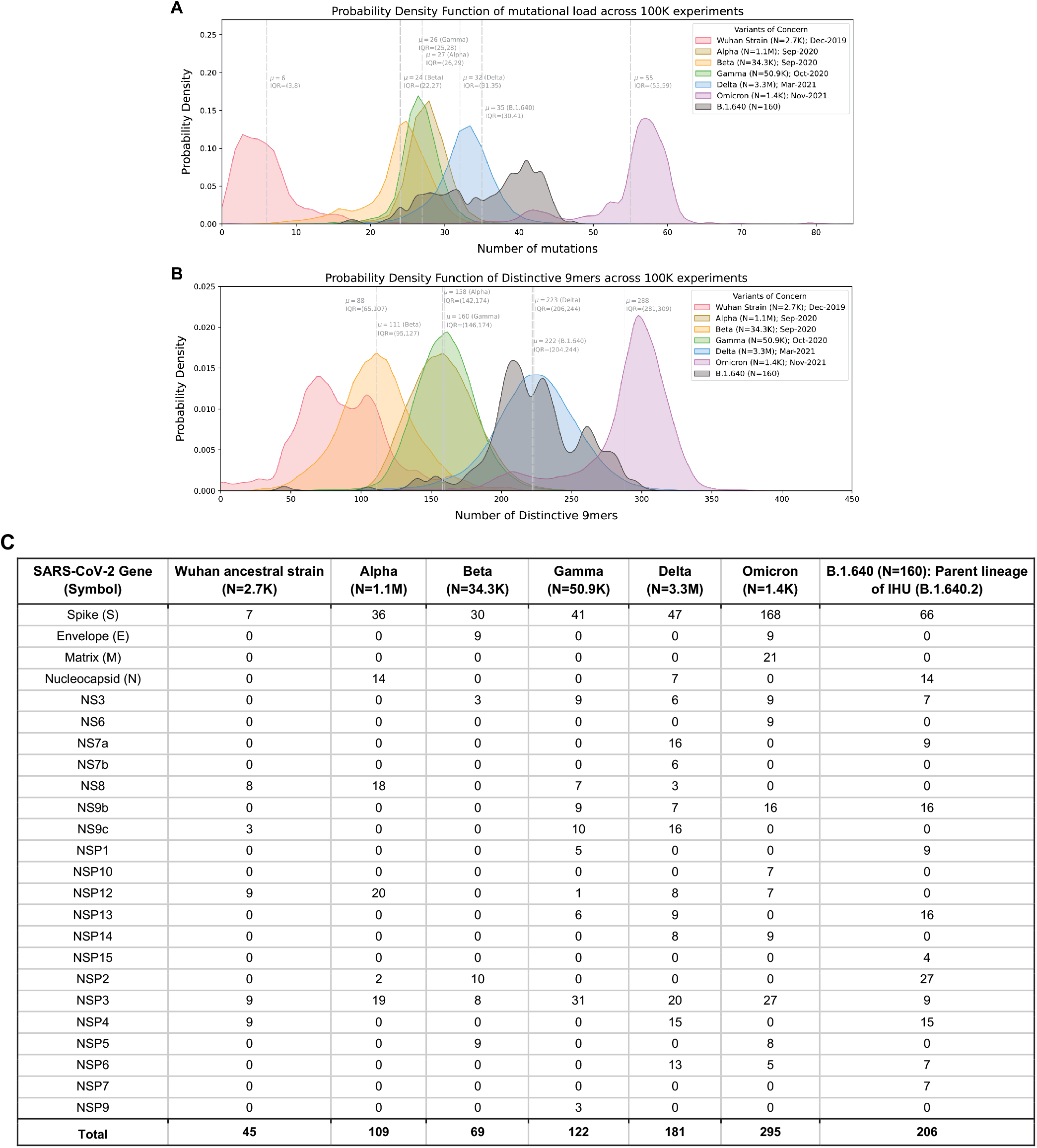
Characterization of the IHU variant (B.1.640.2) containing lineage (B.1.640) shows IHU has more mutational load but similar genomic distinctiveness score when compared with Delta. **(A)** Density plots showing the distribution of the number of mutations observed in SARS-CoV-2 genomes assigned to each listed lineage in the GISAID database. **(B)** Density plots showing 9-mer sequence distinctiveness for VOCs, as measured by the number of distinct 9-mer polynucleotide sequences. **(C)** Summary of the number of 9-mer nucleotides that are distinctive in at least 50,000 among 100,000 experiments for each gene-lineage combination.

The gene-level summary of the distinctive 9-mers across the original SARS-CoV-2 strain, the VOCs, and the B.1.640 lineage are provided (**Figure 3C**). This shows that considering only the spike (S) protein is insufficient to distinguish distinctiveness of the VOCs in a way consistent with their emergence and dominance. For example, while the Delta variant is a higher genome-wide distinctiveness than Alpha, Beta, and Gamma, the distinctiveness values for their respective Spike protein-encoding nucleotide sequences are similar. Further, it shows that there are several distinctive polynucleotides harbored in highly mutated variants that have not yet circulated globally, such as the B.1.640 lineage.

We also computed pairwise phylogenetic distances between all VOCs using four standard metrics (Tajima-Nei,^20^ Jukes Cantor,^21^ Tamura,^22^ and Kimura^23^). All metrics other than Jukes Cantor highlight Gamma as most phylogenetically from Alpha and Beta, while Jukes Cantor more strongly reveals the distance between Delta and all other lineages (**Figure 2D-G**). Notably, none of these phylogenetic distance metrics reveal the strong distinction between Omicron and other lineages which is captured by both mutational load and our genomic distinctiveness metric.

### The emergence of new VOCs has had minimal impact on the genomic distinctiveness of prior VOCs

New variants can emerge that share genomic features with a given prior variant (e.g., due to convergent evolution). In these cases, nucleotide n-mers that were previously considered distinctive for that variant will no longer be considered as such, resulting in the potential tendency of DN9s for any lineage to decrease over time after its emergence. In theory, this could at least partially explain the previously described correlation between time of VOC emergence and genomic distinctiveness. To determine whether this is the case, we performed a modified experiment in which we assessed the impact of Delta on the genomic distinctiveness of Alpha, Beta, and Gamma with respect to the original strain.

Even when comparing only Alpha, Beta, and Gamma genomes deposited before June 2021, the DN9s were similar between these three VOCs (**Figure 4A**). When additionally considering the Delta variant and expanding to include genomes deposited before November 2021, there was only a slight leftward shift in the DN9 distributions for Alpha, Beta, and Gamma (**Figure 4B**). As expected, Delta genomes tended to have substantially more DN9s than the other VOCs (**Figure 4B**). Comparison of this data to the initial analysis (**Figure 1C**) also confirms that the emergence of Omicron did not result in a pronounced leftward shift of the genomic distinctiveness of Delta or the other VOCs. Together, this analysis suggests that not only are newly evolving SARS-CoV-2 variants more genomically distinctive compared to the original strain than prior variants, but they also explore unique nucleotide sequences to generate this distinctiveness.

**Figure 4.**
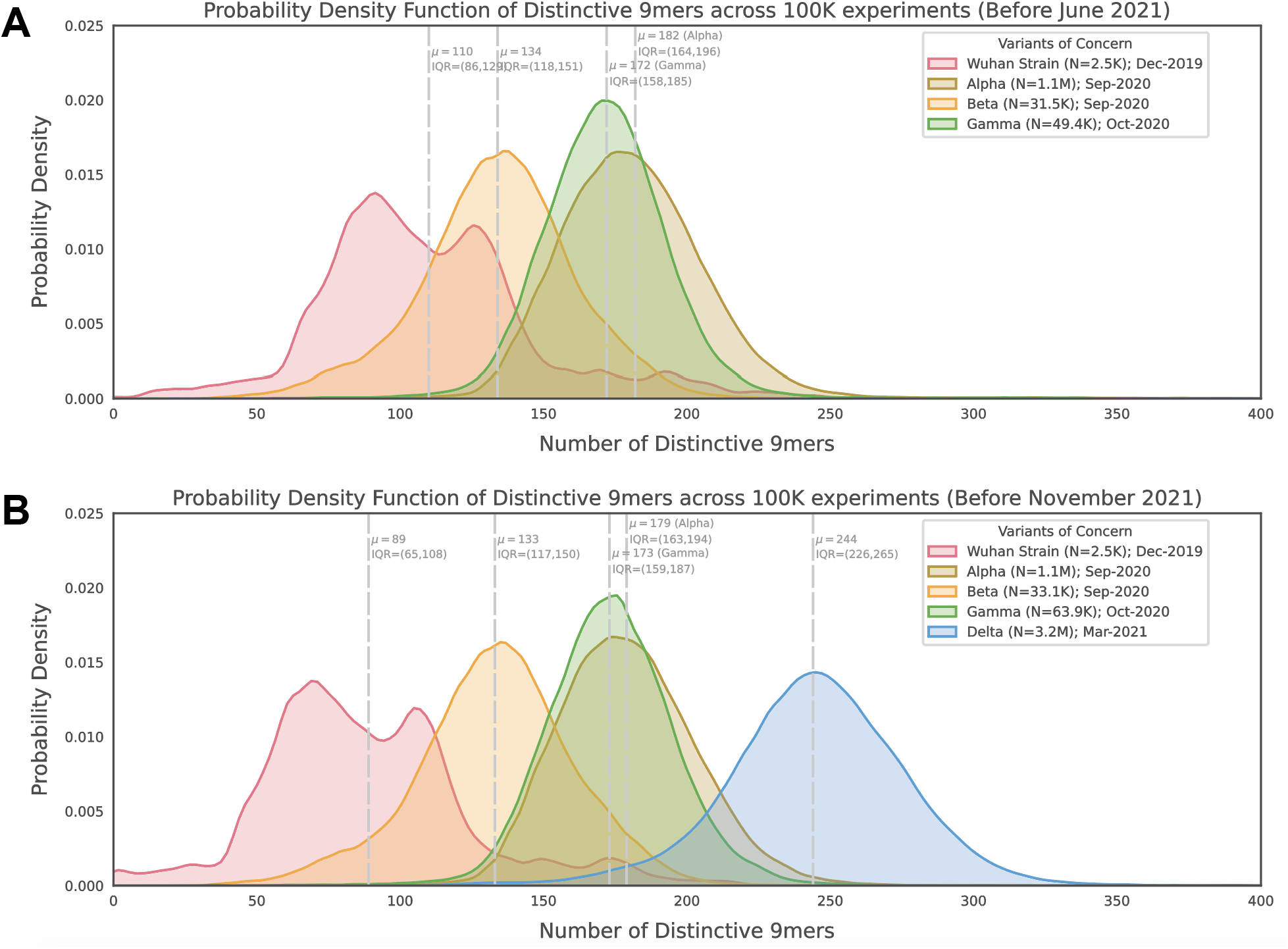
Assessment of the impact of new VOCs on the genomic distinctiveness of prior VOCs. **(A)** Distribution of the number of distinctive 9-mer polynucleotides contained in reference, Alpha, Beta, and Gamma genomes collected before June 2021. Genomes from other VOCs (including Delta) did not contribute at all to this analysis. **(B)** Distribution of the number of distinctive 9-mer polynucleotides contained in reference, Alpha, Beta, Gamma, and Delta genomes before June 2021. Only the VOCs shown in the plot (i.e. not Omicron) contributed to this analysis. Abbreviations: VOCs - variants of concern.

### Increasing distinctiveness within the Delta lineage over time was recently outpaced by the highly distinctive Omicron variant

It is also possible that the correlation between genomic distinctiveness and time of emergence simply reflects an evolutionary clock, i.e. the amount of time between the first detection of SARS-CoV-2 and the emergence of each variant. If this is true, then we would expect to observe the following: (i) genomic distinctiveness within a lineage should increase over time; (ii) genomic distinctiveness between lineages should be similar at any given point in time. To test this hypothesis, we modified our initial analysis to compute polynucleotide distinctiveness distributions for Delta genomes divided into three non-overlapping intervals based on their collection date (April, July, or December 2021).

The distribution of Delta genome DN9s indeed shifted right over time (April: mean 203, IQR 194-218; July: mean 223, IQR 210-239; December: mean 259, IQR 243-276) (**Figure 5**). This indicates that distinctiveness can indeed increase within a lineage with the passage of evolutionary time. However, it is interesting to note that Omicron genomes do tend to have more DN9s than even contemporaneous Delta genomes (i.e. collected in December) (**Figure 5**). This suggests that Omicron may be more distinctive than would be expected from evolutionary time alone.

**Figure 5.**
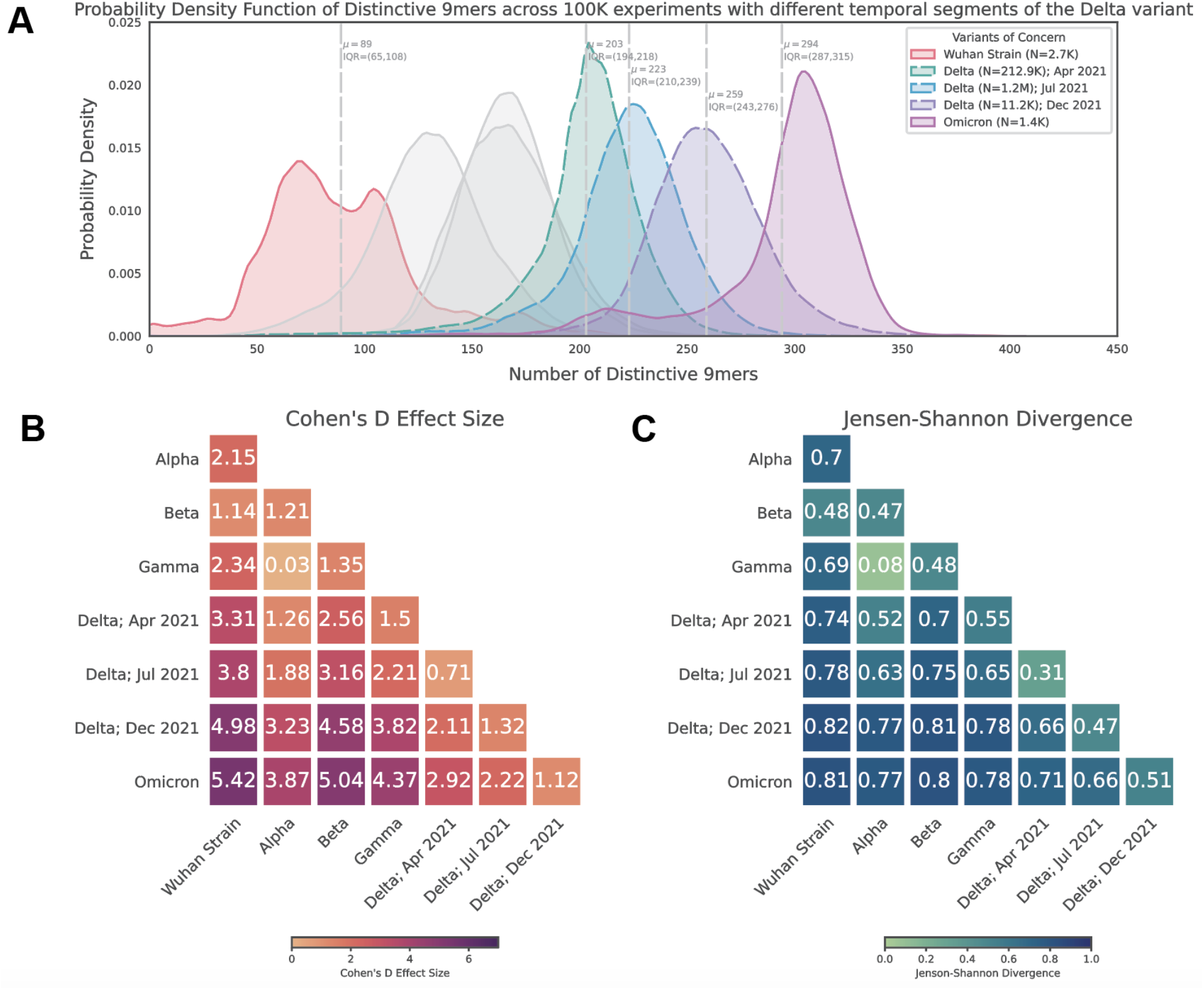
Genomic distinctiveness increases within the Delta lineage over time. **(A)** Histogram showing the distribution of distinctive 9-mer counts for all VOCs, with Delta genomes divided temporally into three groups: early (collected during April 2021), middle (collected during July 2021), and late (collected during December 2021). The distributions shown for the other variants are identical to those shown in Figure 1C, and the Alpha, Beta, and Gamma distributions are shown here in gray (not included in panel legend). Note that each sub-analysis of temporally restricted Delta sequences was performed independently such that in any round (experiment), only one Delta genome was compared to the genomes of other VOCs. That is, early, middle, and late Delta sequences were not compared to each other during the derivation of their distinctive 9-mers. **(B-C)** Pairwise Cohen’s D and Jensen-Shannon Divergence values between the distinctive 9-mer distributions shown in (A). Abbreviations: μ - mean; IQR - interquartile range; VOCs - variants of concern.

### Polynucleotide diversification increases across SARS-CoV-2 lineages over time

In order to compare all SARS-CoV-2 lineages reported to date (rather than restricting to VOCs), we defined another related metric of polynucleotide distinctiveness, termed *A*(1-B)*, which captures the average specificity to a given lineage of all unique n-mers present in that lineage (see **Methods**). This metric correlates with the date at which a lineage first emerged (**Figure 6**), supporting that polynucleotide distinctiveness captures evolutionary drift from the root lineage (i.e., later descendants have less and less in common with each other). Furthermore, this metric places most VOCs (Alpha, Beta, Gamma, Delta, and Omicron) as outliers compared with contemporarily emerging lineages (**Figure 6**). Importantly, this appears to be true even when only considering genomes deposited before or shortly after the emergence of a given VOC (**Figure S3A**), suggesting that this metric could be useful in future predictive analyses of newly identified variants.

**Figure 6.**
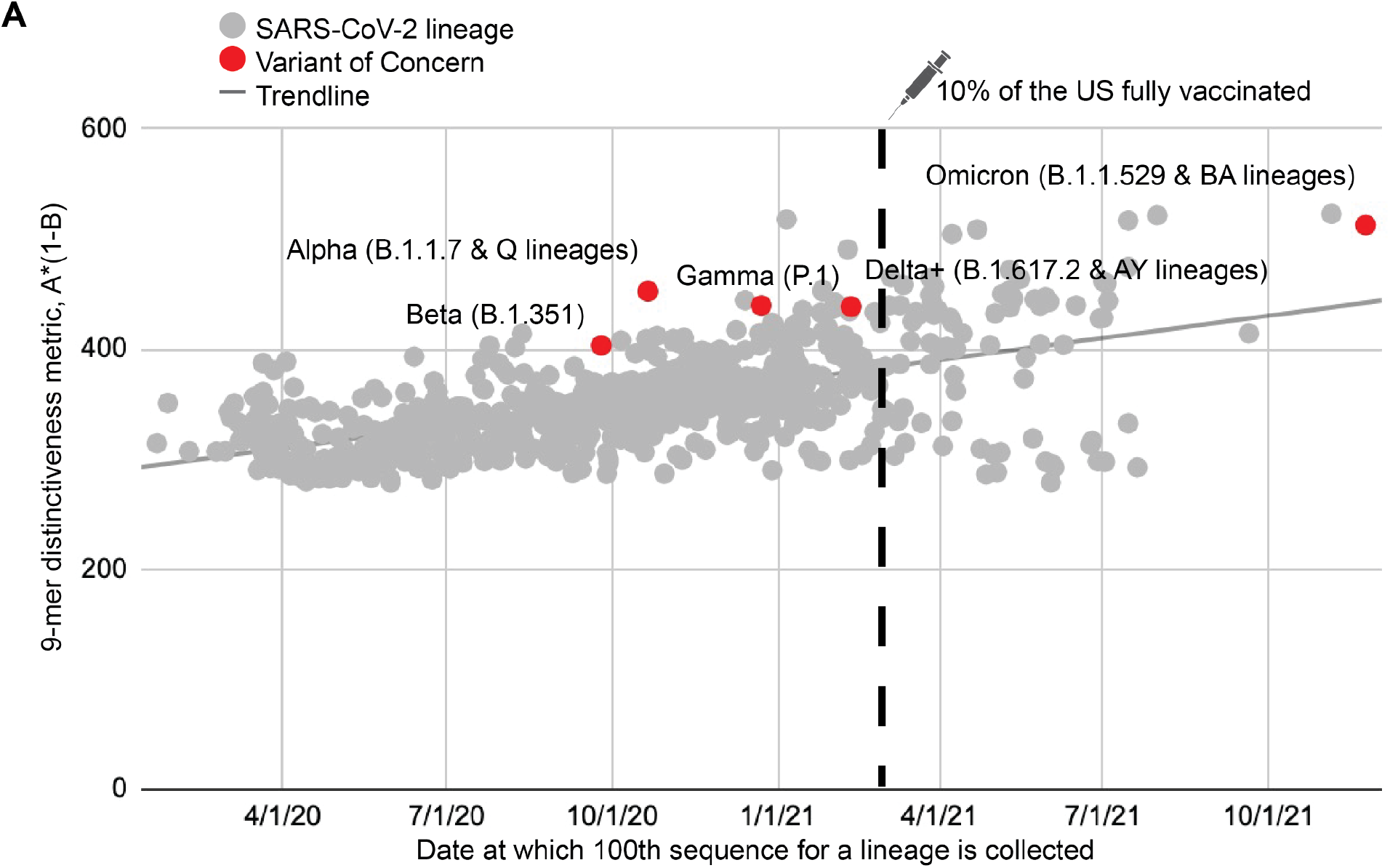
Temporal trends in polynucleotide sequence diversity. Global metric for 9-mer distinctiveness plotted against lineage emergence date for the 1,363 GISAID lineages with over 200 sequences reported. Variants of concern are labeled and highlighted in red. The date when 10% of the United States population was fully vaccinated is highlighted with a syringe symbol and a vertical dotted line.

## Discussion

### Summary of results

The analyses described here demonstrate that SARS-CoV-2 has continually evolved into variants with increasing levels of genomic distinctiveness with respect to the original strain. Among the lineages that have been classified as VOCs and spread widely, the recently emerged and rapidly spreading Omicron variant has both the highest number of amino acid mutations and the highest distinctiveness at the polynucleotide level relative to other VOCs. While it is generally expected that any virus will accumulate more mutations over time, it appears that the distinctiveness of VOCs at the polynucleotide level provides additional context beyond the number of mutations acquired. This investigation of the diversity of the nucleotide language explored by SARS-CoV-2 can be considered as a supplement to existing phylogenetic classifications, which themselves were crucial to conduct the presented analyses. Through our publicly available interface (https://academia.nferx.com/GENI), this metric will be continually applied as new variants emerge to understand the degree of distinctiveness for any new lineages with respect to prior or current VOCs.

### Implications and future directions

The fact that Delta sequences collected during December 2021 are more distinctive with respect to the original strain than Delta sequences from April or July 2021 indicates that this metric is at least partially reflective of evolutionary time. This finding is expected given the known evolution of Delta sub-lineages with additional mutations compared to the original VOC.^24^ However, the increased distinctiveness of Omicron sequences compared to contemporaneous Delta sequences (i.e., collected in December) interestingly suggests that Omicron may be more distinctive than would be expected from evolutionary time alone. Thus, sequential surges of COVID-19 can be driven by increasingly distinctive VOCs which continue evolving toward more distinctive lineages during their circulation and are ultimately supplanted by a lineage which is ahead of schedule in terms of genomic distinctiveness. This is consistent with the previous suggestion that VOCs can emerge because of saltational events that occur over extended incubation times in immunocompromised individuals who clear SARS-CoV-2 less efficiently.^25–27^

The ability of this metric to predict viral fitness and which variants are most likely to outcompete others needs to be further investigated. Currently, the likelihood that a new lineage will cause surges is speculated based on the number and constellation of mutations that it harbors, particularly based on the Spike protein’s mutations. For example, because the Omicron variant was so heavily mutated as compared to prior strains, including mutations in regions of the Spike protein known to be functionally important (e.g., RBD, furin cleavage site, NTD), it was quickly hypothesized that Omicron would drive surges widely. Given that the propensity of an emerging variant to drive surges is related to several factors including immune evasive capacity,^3^ we hypothesize that variants which are more distinctive compared to lineages that have previously circulated or are contemporaneously circulating in the same region may be likely candidates for new VOCs. This hypothesis is consistent with the observation that the highly distinctive Omicron variant has already overtaken Delta as the dominant variant in many regions of the world as of January 5, 2022, whereas the highly mutated but less distinctive IHU variant (which emerged before Omicron) was apparently unable to outcompete Delta. It should be emphasized that the IHU variant has several distinctive polynucleotide n-mers that are not observed in previous VOCs which circulated significantly across the globe (**Figure 3C**). It will be interesting to determine whether such n-mers that are distinctive to a non-dominant but highly mutated variant are “recycled” by VOCs that arise in the future. One benefit of our distinctiveness metric is that even a single sequence of a potential new lineage can be rapidly assessed for distinctiveness relative to prior VOCs or time- and geography-matched SARS-CoV-2 genomes, with further statistical confidence gained as more sequences from such a new lineage are deposited. The free availability of our software (GENI) on the internet should further aid in near-real-time evaluation of such new lineages or sub-lineages.

There are several other questions to which this method can be applied, some of which we have explored preliminarily. For example, we have evaluated genomic distinctiveness between lineages that have previously been or are currently labeled as variants of interest (VOIs) rather than VOCs.^3^ Here we do not observe any Delta-like or Omicron-like outliers with respect to their polynucleotide distinctiveness (**Figure S2A**), which is consistent with the fact that these lineages have not been widely responsible for large COVID-19 surges. As a control, we also confirmed that seasonal human-infecting coronavirus (HCoV) genomes, which are similar in size to SARS-CoV-2 genomes, are substantially more distinct from the original (Wuhan) SARS-CoV-2 strain than any VOCs (**Figure S2B-C**). It should be noted that the two-dimensional (2-D) visualization provided by this analysis (**Figure S2B**) does not imply a transition of VOCs toward a more seasonal coronavirus-like genome, although methods to understand whether such an evolutionary process is likely to occur at some point will be important to develop as part of follow-up research studies. Others have recently reported that infections with Omicron are less likely to cause severe disease than Delta, but these analyses are limited by small sample sizes and confounding variables such as vaccination status (including time since vaccination and booster doses) and higher rates of Omicron infections among young individuals.^28–30^

### Methodological considerations and study limitations

Most of the analyses presented here refer to 9-mer polynucleotides, but it is reasonable to consider polynucleotides of various lengths. Indeed, we found that this metric was robust to smaller and larger polynucleotides, with particularly strong separation between some VOCs observed for n-mers between 15 and 30 nucleotides. The presented results based on 9-mers should be considered as an example to illustrate the utility of this metric rather than the definitive resolution. There are also some limitations to these analyses. First, the number of Omicron sequences currently available in the GISAID database is low compared to other VOCs such as Delta. Our protocol, which samples genomes with replacement, could result in oversampling of Omicron sequences. This limitation will be addressed in the coming months as more Omicron sequences are deposited. Second, while we consider all sliding nucleotide 9-mers, it is also worth exploring similar metrics of genomic diversity while constraining to protein-coding nucleotide n-mers or amino acid n-mers themselves. Third, both methods presented here compare one lineage to one or many others, and thus they are sensitive to the lineage composition in the complement group. For example, several Delta sublineages show relatively low polynucleotide distinctiveness through the *A*\*(1-*B*) metric, but this is likely due to the fact that they are being compared to other Delta sub-lineages which are highly related and quite prevalent in the dataset (**Figure S3B**). Finally, further investigation into the relationship between our genomic distinctiveness metrics and other features such as phylogenetic depth and evolutionary time are warranted, although we have performed preliminary sensitivity analyses which suggest that our metrics provide additional value beyond these features.

### Concluding remarks

In summary, we present here a new methodology and an internet resource that will enable researchers to rapidly assess the distinctiveness of new SARS-CoV-2 lineages relative to any prior or contemporary lineages. We encourage further investigation into whether this method can help to classify lineages as VOCs earlier, study how vaccination impacts SARS-CoV-2 genomic diversity, and determine if or when SARS-CoV-2 will transition toward endemicity or seasonality.

## Methods

### Quantification of number of distinct n-mers for SARS-CoV-2 Variants of Concern (VOCs)

The number of distinctive n-mers for SARS-CoV-2 sequences from the original reference strain (PANGO lineage A) and five prior or current VOCs (Alpha, Beta, Gamma, Delta, and Omicron) were calculated using genomes obtained from the GISAID database (https://www.gisaid.org).^6,7^ Specifically, genomes were sampled with replacement for each VOC by mapping to their corresponding PANGO lineages. We generated 100,000 sets of 6 genomes (using a total of 287,739 unique genomes) and then computed the number of unique 9-mer polynucleotides for every combination of the 6 VOCs in each of the 100,000 sets. For a given genome, 9-mer polynucleotides that contain non-ACGT characters were discarded. We refer to this overall process as performing 100,000 experiments. In each experiment, distinctive nucleotide 9-mers (DN9s) for a given variant were defined as those which were present in the genome of that variant but not in the genomes of any other variants. We also determine the distribution of prevalent DN9s across different SARS-CoV-2 genes. Here, DN9s that occur in at least 50,000 experiments (out of 100,000 experiments) are deemed prevalent. This procedure was repeated for n-mer sequences of various lengths, including 3, 6, 9, 12, 15, 18, 21, 24, 30, 45, 60, 75, 120, and 240 nucleotides.

For each pair of variants, we computed the Cohen’s D between the distributions of distinctive n-mer sequence counts using the following equation:

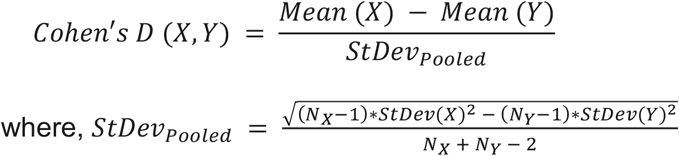

The Jensen-Shannon (JS) Divergence values were computed using the SciPy package (version 1.7.3) in Python (version 3.7.10).

For all analyses, only the genomes which were annotated as high-quality and complete in GISAID were considered. As expected, this resulted in a very tight distribution of sequence length and total number of sliding n-mers per SARS-CoV-2 sequence (**Figure S4**).

### Mutational Load of SARS-CoV-2 VOCs

For sequences obtained from GISAID, we determined the number of amino-acid mutations (insertions, substitutions, and deletions as reported in GISAID) relative to the originally deposited Wuhan-Hu-1 strain of SARS-CoV-2 (GenBank accession number MN908947: https://www.ncbi.nlm.nih.gov/nuccore/MN908947.3/).^18^ We generated a distribution of the number of mutations for each VOC and then performed pairwise comparisons between all variants by calculating Cohen’s D and JS divergence.

### Phylogenetic reconstruction of SARS-CoV-2 VOCs

The multiple sequence alignment (MSA) of sub-sampled SARS-CoV-2 genomes (3,095 genomes randomly sampled from ‘*Open data’*) was derived from the Nextstrain database (https://nextstrain.org/), accessed on 20th December, 2021.^31^ The MSA was further processed to retain 1539 sequences belonging to the strains of interest: Alpha (97 genomes), Beta (20 genomes), Gamma (28 genomes), Delta (1,380 genomes), Omicron (2 genomes) and PANGO lineage A (12 sequences) (**Table S1**). The pruned MSA was then used to calculate the phylogenetic distance using four established metrics: (1) Tajima-Nei,^20^ (2) Tamura,^22^ (3) Jukes Cantor,^21^ and (4) Kimura.^23^ The pairwise phylogenetic distance between any two VOCs was taken as the mean of the phylogenetic distances between all pairs of sequences belonging to those variants.

### Calculation of the alternative n-mer distinctiveness metric A*(1-B) that incorporates intra-lineage conservation scores

To compare the distinctiveness of each SARS-CoV-2 lineage versus all other lineages in GISAID, we defined a related metric to capture the distinctiveness of n-mers for a specific SARS-CoV-2. For a given lineage, *l*, we calculate the following n-mer distinctiveness metric:

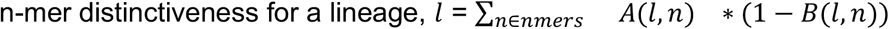

where *A*(*l, n*) is the fraction of sequences in GISAID of lineage l that contain a specific n-mer, *n*, and *B*(*l, n*) is the fraction of sequences in GISAID not of lineage *l* that contain n-mer *n*. The sum is over all n-mers that are found for lineage *l*. Results presented here specifically use 9 as the n-mer size.

### Data sources

All SARS-CoV-2 sequences were downloaded from GISAID (https://www.gisaid.org/). The 2021-12-13 version of the raw FASTA file and supporting metadata were used.

## Data Availability

All data produced are available online at https://academia.nferx.com/GENI

## Supplementary Material

**Figure S1.**
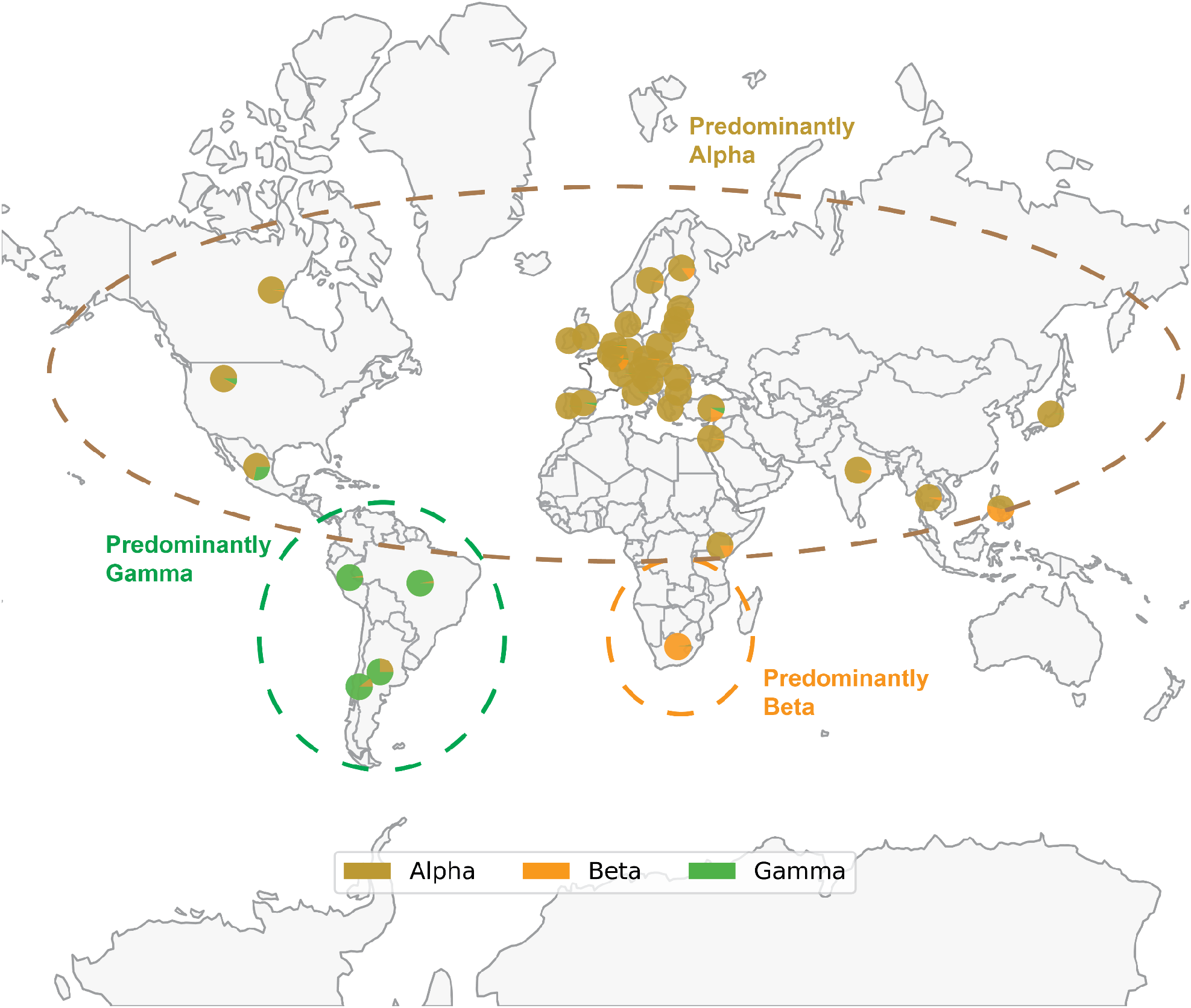
Map of SARS-CoV-2 VOC prevalence by geographic region. Geographical distribution of Alpha (B.1.1.7), Beta (B.1.351) and Gamma (P.1) variants based on sequences deposited in GISAID through December 14, 2021. Each pie chart shows the proportion of Alpha, Beta or Gamma sequences deposited in the country. Note that the denominator is the number of sequences labeled as any of these three variants, rather than the total number of sequences deposited in that country. Thus, each pie chart answers the following question: “Of all genomes deposited in a given country which were assigned as Alpha, Beta, or Gamma, what proportion of genomes was assigned to each of these three lineages?” The prevalence of Delta and Omicron are not shown to better highlight the geographical distribution of Alpha, Beta, and Gamma; however, Delta and Omicron are currently or have previously been highly prevalent in the regions shown. Only countries where at least 1000 sequences are deposited are shown. The variants depicted, which circulated at approximately the same time, generally became prominent in geographically distinct regions.

**Figure S2.**
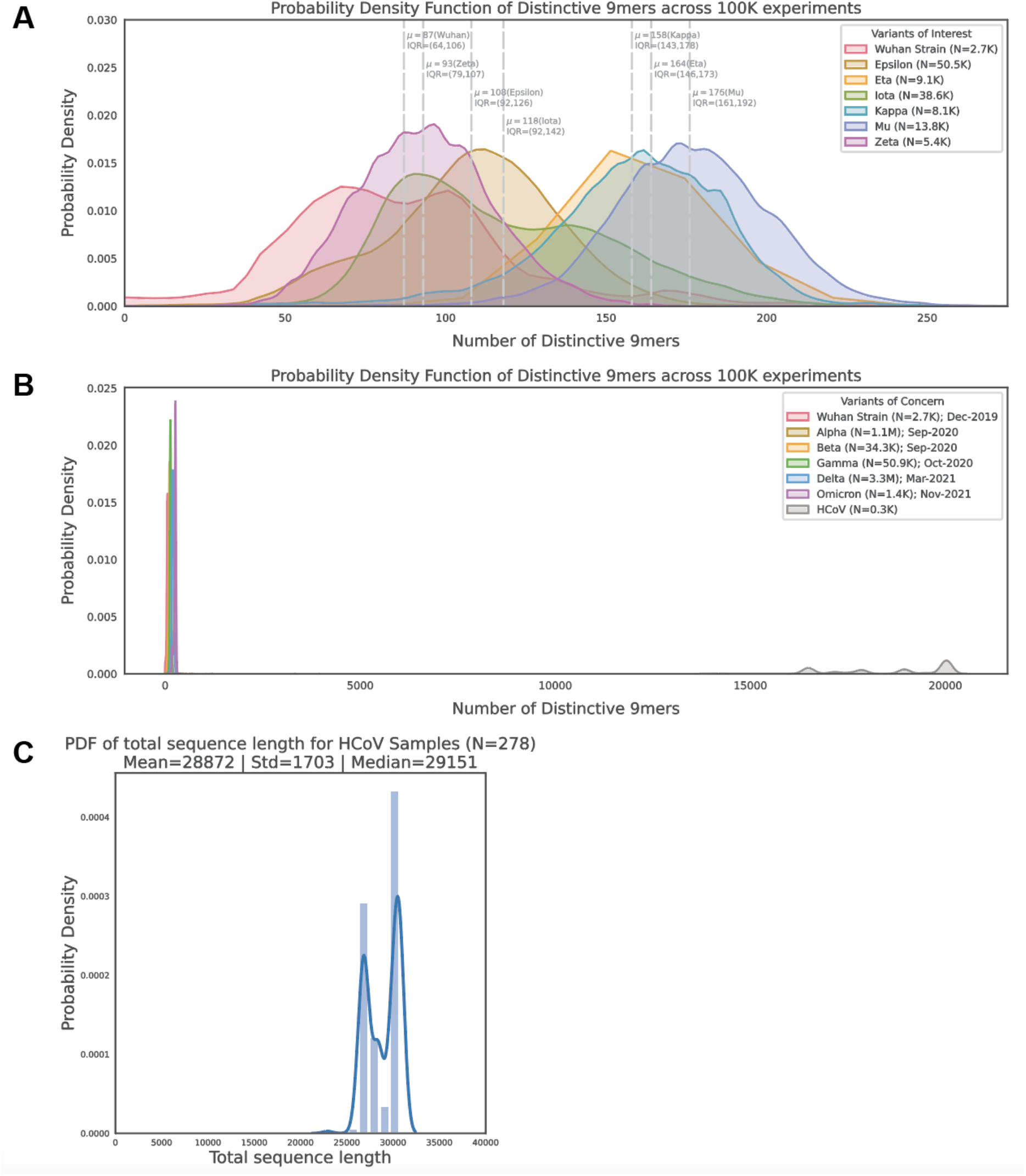
Polynucleotide distinctiveness analysis for various alternate sequence sets. **(A)** Distribution of the number of distinctive 9-mers for SARS-CoV-2 variants of interest. **(B)** Distribution of the number of distinctive 9-mers for SARS-CoV-2 variants of concern and human-infecting seasonal coronavirus (HCoV) genomes, showing that other virus species with similarly sized genomes are significantly more distinctive from the original SARS-CoV-2 strain than any of the SARS-CoV-2 VOCs considered. **(C)** Distribution of the genome length, in nucleotides, for the HCoV sequences used in panel (C). The genome length of SARS-CoV-2 is approximately 30 kilobases. Abbreviations: HCoV - human seasonal coronavirus.

**Figure S3.**
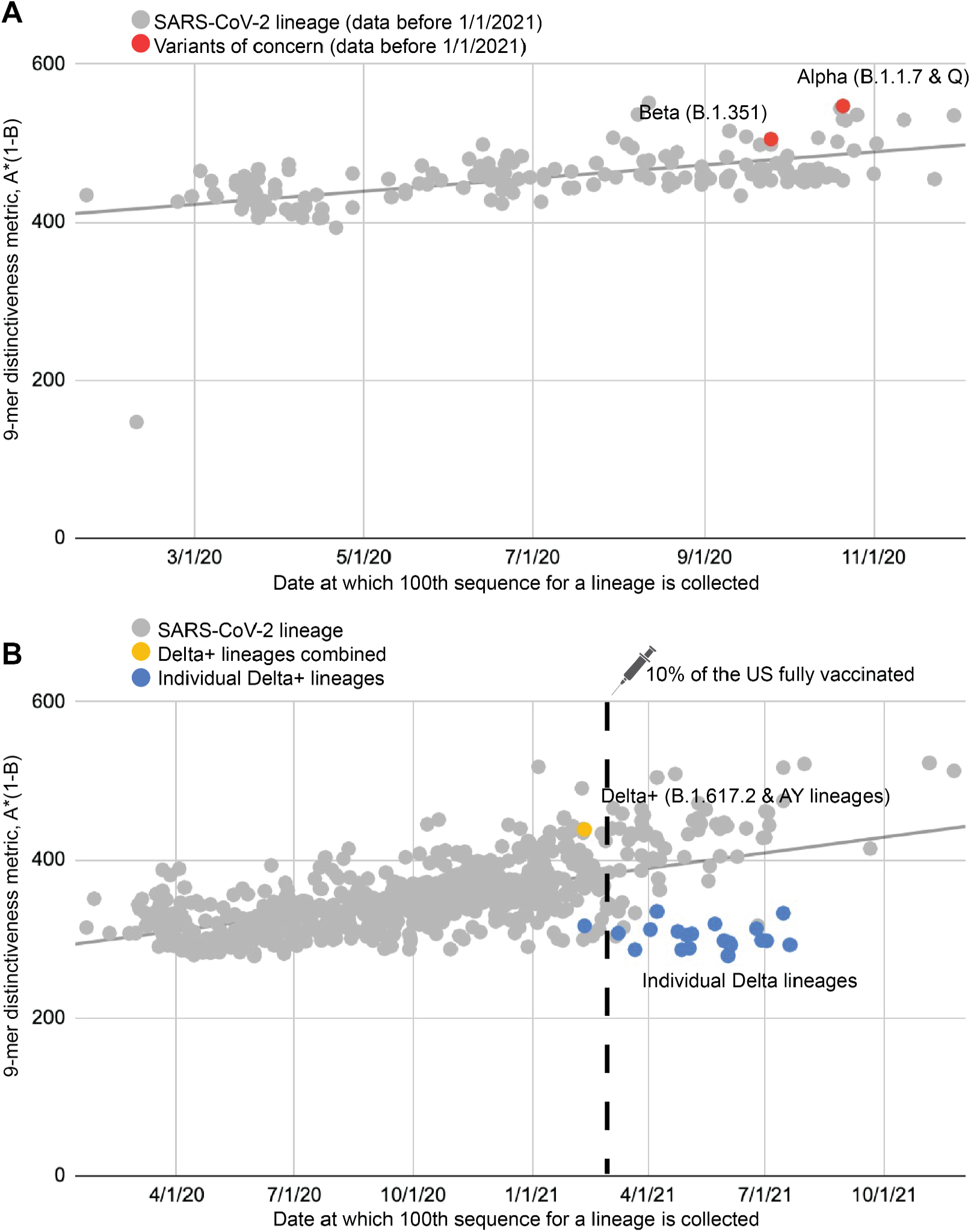
9-mer distinctiveness metric, considering the Delta Plus lineages separately and at an earlier time-point. **(A)** Here we adapt Figure 4A from the main text of this manuscript, but only considering sequences collected prior to January 2021. This indicates that the Alpha variant was highly distinctive as assessed by this metric around its time of emergence. **(B)** Here we adapt Figure 4A from the main text of this manuscript, but with Delta Plus lineages also considered separately (blue dots).

**Figure S4.**
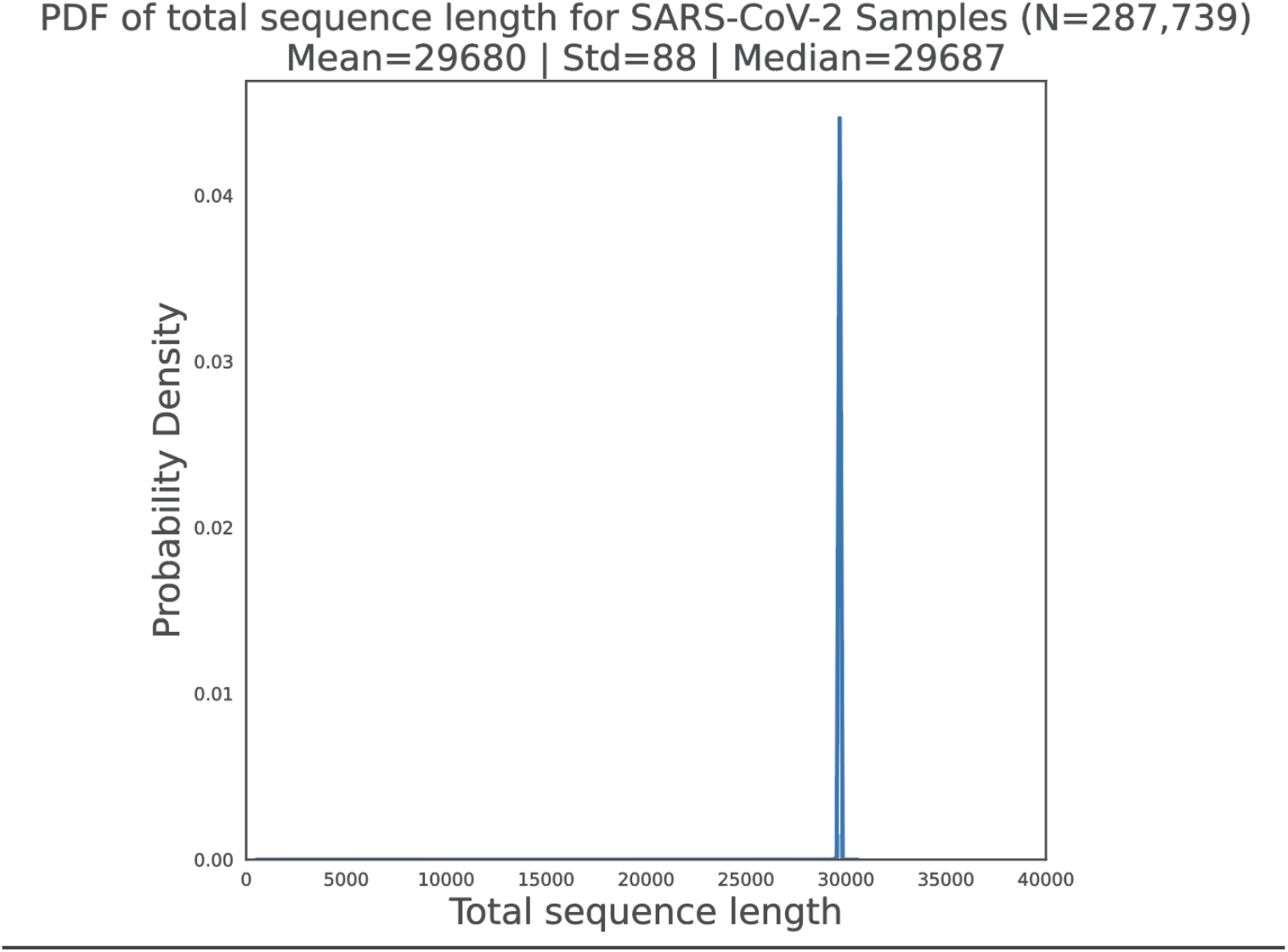
**Distribution of the genome length, in nucleotides, for the SARS-CoV-2 sequences (original strain and VOCs) used throughout this study**.

**Table S1.** Mapping of PANGO lineages to World Health Organization (WHO) nomenclature variant names used throughout the analysis. Source: https://www.cdc.gov/coronavirus/2019-ncov/variants/variant-classifications.html

| **Variant Name** | **PANGO Lineage (\* indicates all descendant lineages)** |
| --- | --- |
| Original Strain | A |
| Alpha | B.1.1.7, Q* |
| Beta | B.1.351* |
| Gamma | P.1* |
| Delta | B.1.617.2, AY* |
| Omicron | B.1.1.529, BA* |
| Epsilon | B.1.427, B.1.429 |
| Eta | B.1.525 |
| Iota | B.1.526 |
| Kappa | B.1.617.1 |
| Mu | B.1.621, B.1.621.1 |
| Zeta | P.2 |

**Table S2.** Cohen’s D values comparing the distributions of distinctive nucleotide 9-mer counts in each VOC versus the original SARS-CoV-2 strain for various n-mer lengths. The highest Cohen’s D value for each variant is highlighted.

| n-mer length | Alpha | Beta | Gamma | Delta | Omicron |
| --- | --- | --- | --- | --- | --- |
| 3 | 0 | 0 | 0 | 0 | 0 |
| 6 | 0.53 | 1.5 | 3.11 | 1.4 | 4.49 |
| 9 | 2.15 | 1.14 | 2.34 | 4.04 | 5.42 |
| 12 | 2.9 | 1.56 | 3.26 | 4.4 | 5.95 |
| 15 | 3.04 | 1.58 | 3.38 | 4.43 | 6 |
| 18 | 3.09 | 1.58 | 3.41 | 4.43 | 6.03 |
| 21 | 3.13 | 1.56 | 3.4 | 4.46 | 6.03 |
| 24 | 3.17 | 1.54 | 3.4 | 4.46 | 6.01 |
| 30 | 3.22 | 1.51 | 3.4 | 4.46 | 5.95 |
| 45 | 3.29 | 1.4 | 3.3 | 4.39 | 5.68 |
| 60 | 3.32 | 1.28 | 3.18 | 4.27 | 5.29 |
| 75 | 3.29 | 1.19 | 3.06 | 4.15 | 4.95 |
| 120 | 3.06 | 0.96 | 2.71 | 3.77 | 4.32 |
| 240 | 2.63 | 0.58 | 2.27 | 3.06 | 2.97 |

